# The 2025 Los Angeles Wildfires and Outpatient Acute Healthcare Utilization

**DOI:** 10.1101/2025.03.13.25323617

**Authors:** Joan A. Casey, Yuqian M. Gu, Lara Schwarz, Timothy B. Frankland, Lauren B. Wilner, Heather McBrien, Nina M. Flores, Arnab K. Dey, Gina S. Lee, Chen Chen, Tarik Benmarhnia, Sara Y. Tartof

## Abstract

January 2025 brought devastating wildfires to Los Angeles (LA) County, California, causing poor air quality, destroying homes and businesses, and displacing thousands of people. We used electronic health record data from 3.7 million Kaiser Permanente Southern California members to promptly determine if the 2025 LA Fires increased outpatient acute healthcare utilization. We created exposure categories using the maximum wildfire burn zone reached by an LA or Ventura County wildfire as of January 16, 2025. Highly-exposed members resided in census tracts located <20km from burn zones and moderately-exposed members lived in tracts ≥20km but within LA County. We identified daily outpatient and virtual acute care visits in five categories: all-cause, cardiovascular, injury, neuropsychiatric, and respiratory. We conducted 2-stage interrupted time-series analyses using machine-learning algorithms to determine if and by how much the 2025 LA Fires increased acute healthcare utilization. Across the week following the January 7 LA Fires ignitions, virtual respiratory visits were 41% (95% empirical confidence interval [eCI]: 26%, 56%) higher and 34% (95% eCI: 17%, 52%) higher than expected in highly- and moderately-exposed groups, respectively, totaling 3,221 excess visits. Similarly, both exposure groups had approximately 35% more virtual cardiovascular visits than expected over the same period. Among highly-exposed members, outpatient and virtual injury visits and outpatient neuropsychiatric visits were ≥18% higher than expected on January 7. Substantial increases in acute healthcare utilization driven primarily by virtual care-seeking were observed following the LA Fires. As disruptive climate events increase, such data are essential to inform healthcare preparedness and response.

**Significance statement:** Climate hazards like the 2025 LA Fires will increasingly impact US cities. We identified increased acute outpatient and virtual care use for cardiovascular, injury, neuropsychiatric, and respiratory conditions over the week post-LA Fires ignitions from Kaiser Permanente members living <20 km from a burn zone or anywhere in LA County. This included an excess of 2,424 (+35%) cardiovascular and 3,221 (+35%) respiratory virtual visits. By applying our estimates to all LA County residents, we estimated 16,171 excess cardiovascular and 21,541 excess respiratory virtual care visits occurred during the week following ignition. We identified increased acute care visits, primarily driven by virtual care-seeking, suggesting this healthcare service should be expanded during disruptive climate events.

## Introduction

On January 7, 2025, the Palisades and Eaton fires ignited in Los Angeles County (LA), California, followed by other fires in LA and neighboring Ventura County. Fueled by climate change and strong Santa Ana winds, the fires burned >38,000 acres and destroyed >16,000 structures by January 19, 2025. Officials identified 29 direct fatalities,^1^ but the true health burden will exceed that tally. Smoke caused fine particulate matter (PM_2.5_) concentrations to exceed the U.S. Environmental Protection Agency 24-hour standard of 35 *μ*g/m^3^ for several days and to briefly top 400 *μ*g/m^3^ in LA on January 8 before returning to background levels by January 12. ^2^

Previous studies have identified short-term wildfire PM_2.5_ as a risk factor for respiratory acute care visits and all-cause mortality,^3^ and wildfire burn zone proximity as a risk factor for adverse mental health outcomes, injuries, and mortality.^4,5^

Here, we rapidly assess the January 2025 LA Fires’ impact on acute outpatient healthcare utilization at Kaiser Permanente Southern California (KPSC).

## Methods

KPSC is an integrated healthcare system serving more than 4.7 million individuals, and its electronic health record (EHR) data contain longitudinal records of patient residential addresses, sociodemographic characteristics, and diagnoses across all care settings. This study included all KPSC members from November to January 2022–2025, allowing 30-day enrollment gaps (**eFigure 1**). We did apply enrollment criteria for children <3 years old.

We classified census tracts *a priori* into three exposure levels based on their proximity to the maximum burn zone of seven LA-area wildfires as of January 17, 2025: Palisades, Eaton, Kenneth, Hurst, Lidia, Auto, and Sunset.^1^ Highly exposed members resided in census tracts located <20km from a wildfire burn zone and moderately exposed members lived in tracts ≥20km away but within LA County.

We identified daily virtual and outpatient visits in five disease categories using *International Classification of Diseases, Tenth Revision* codes (all-cause, cardiovascular [I00-I99], injury [S00-T88], neuropsychiatric [F01-F99], and respiratory [J00-J99]). We aggregated visits by type, cause, day, and wildfire exposure category.

Time-varying covariates included daily maximum and minimum temperature and humidity, wind velocity, and surface downward shortwave radiation from gridMET and weekly wastewater surveillance data on levels of three respiratory viruses: flu, respiratory syncytial virus (RSV), and SARS-CoV-2.^6^

We used a novel 2-stage interrupted time-series (ITS) analysis to assess the relationship between the LA Fires and daily outpatient acute care utilization in the week (primary) and two weeks (secondary) following the January 7 LA Fires ignitions among highly and moderately exposed groups. Briefly, this 2-stage ITS analysis was coupled with hybrid machine learning algorithms (e.g., Prophet-Extreme Gradient Boosting) to optimize the prediction of counterfactual trends (acute care utilization in the absence of the LA Fires), and the optimal parameter set was determined by minimizing the root mean square error across cross-validation folds (**eMethods**). This approach may reduce bias present in traditional estimates.^7^ We used a Monte Carlo simulation approach including 1,000 model iterations to estimate 95% empirical confidence intervals.

In a sensitivity analysis, we assessed changes in respiratory visits among minimally exposed members who lived in tracts ≥20km from wildfire burn zones in non-LA KPSC catchment counties (Imperial, Kern, Orange, Riverside, San Bernardino, San Diego, San Luis Obispo, Santa Barbara, and Ventura).

The study protocol was approved by the KPSC and WCG Institutional Review Boards who waived the requirement for informed consent. Analyses were conducted using R Statistical Software version 4.4.1 and Python version 3.12.2. Relevant code is publicly available via GitHub (https://github.com/heathermcb/los_angeles_2025_fire_disasters).

## Results

Our study population consisted of 3.7 million KPSC members of all ages (**Table 1**), of whom 305,258 were highly exposed (**Figure 1**), accounting for nearly 13% of all LA residents living within <20km of an LA Fire burn zone.

**Table 1.** Characteristics of the Kaiser Permanente Southern California study population, Nov 2022-Jan 2025.

|  | Exposure |  |  |  |
| --- | --- | --- | --- | --- |
|  | Total | Highly exposed<sup>a</sup> | Moderately exposed<sup>a</sup> | Minimally exposed<sup>a</sup> |
|  | n=3,717,210 | n=305,258 | n=1,373,419 | n=2,038,533 |
| **Age, years<sup>b</sup>** |  |  |  |  |
| Median (IQR) | 42 (21, 62) | 46 (26, 65) | 42 (22, 61) | 42 (21, 61) |
| **Sex, n (%)** |  |  |  |  |
| Female | 1941970 (52.2%) | 158011 (51.8%) | 723153 (52.7%) | 1060806 (52.0%) |
| Male | 1774871 (47.7%) | 147202 (48.2%) | 650138 (47.3%) | 977531 (48.0%) |
| Other | 167 (<0.1%) | 22 (<0.1%) | 68 (<0.1%) | 77 (0.0%) |
| Unknown | 202 (<0.1%) | 23 (<0.1%) | 60 (<0.1%) | 119 (0.0%) |
| **Race and ethnicity<sup>c</sup>, n (%)** |  |  |  |  |
| Hispanic | 1627406 (43.8%) | 98493 (32.3%) | 719131 (52.4%) | 809782 (39.7%) |
| Non-Hispanic Asian | 426054 (11.5%) | 118389 (38.8%) | 160361 (11.7%) | 221815 (10.9%) |
| Non-Hispanic Black | 290701 (7.8%) | 43878 (14.4%) | 158081 (11.5%) | 117106 (5.7%) |
| Non-Hispanic White | 1078707 (29.0%) | 15514 (5.1%) | 231239 (16.8%) | 729079 (35.8%) |
| Unknown | 173378 (4.7%) | 16137 (5.3%) | 60910 (4.4%) | 96331 (4.7%) |
| Other | 120964 (3.3%) | 12847 (4.2%) | 43697 (3.2%) | 64420 (3.2%) |
| IQR, interquartile range<br><sup>a</sup> Based on the maximum wildfire burn zone reached by an LA or Ventura County wildfire as of January 16, 2025, highly exposed members resided in a census tract located <20km, moderately exposed members lived in tracts ≥20km but within LA County, and minimally exposed members lived in tracts ≥20km in other KPSC catchment counties (Imperial, Kern, Orange, Riverside, San Bernardino, San Diego, San Luis Obispo, Santa Barbara, and Ventura).<br><sup>b</sup> Age was calculated as of January 7, 2025 or death.<br><sup>c</sup> Other includes individuals of multiple races, Native American and Alaskan Native, Pacific Islander, and other. |  |  |  |  |

**Figure 1:**
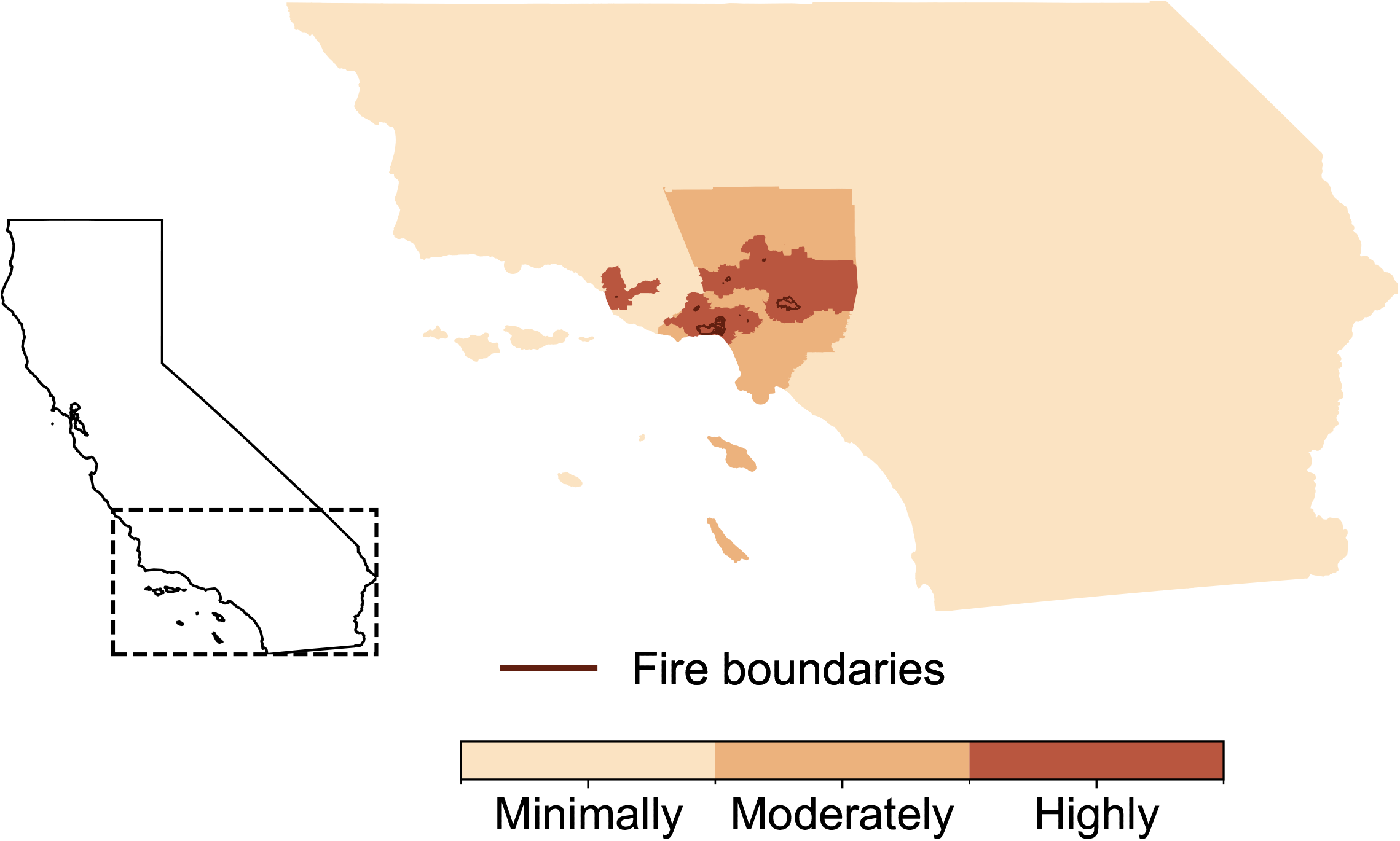
The seven January 2025 Los Angeles (LA) wildfires overlaid with highly, moderately, and minimally exposed census tracts. The three exposure categories included: (1) highly exposed census tracts within 20km of a wildfire burn zone; (2) moderately exposed census tracts >20km from a wildfire burn zone but within LA County; (3) minimally exposed census tracts outside LA County but within the Kaiser Permanente Southern California catchment area. The map depicts the maximum wildfire burn zone reached by the seven LA wildfires as of January 17, 2025.

Over the week following the ignition of the January 7 LA Fires, there were 35% more cardiovascular-related (n=2,424) and respiratory-related (n=3,221) virtual visits than expected in among the combined highly- and moderately-exposed groups (**Figure 2, eFigure 2, eTables 1-8**). By applying these estimates to all LA County residents, we estimated 16,171 excess cardiovascular and 21,541 excess respiratory virtual care visits occurred during the week following ignition.

**Figure 2:**
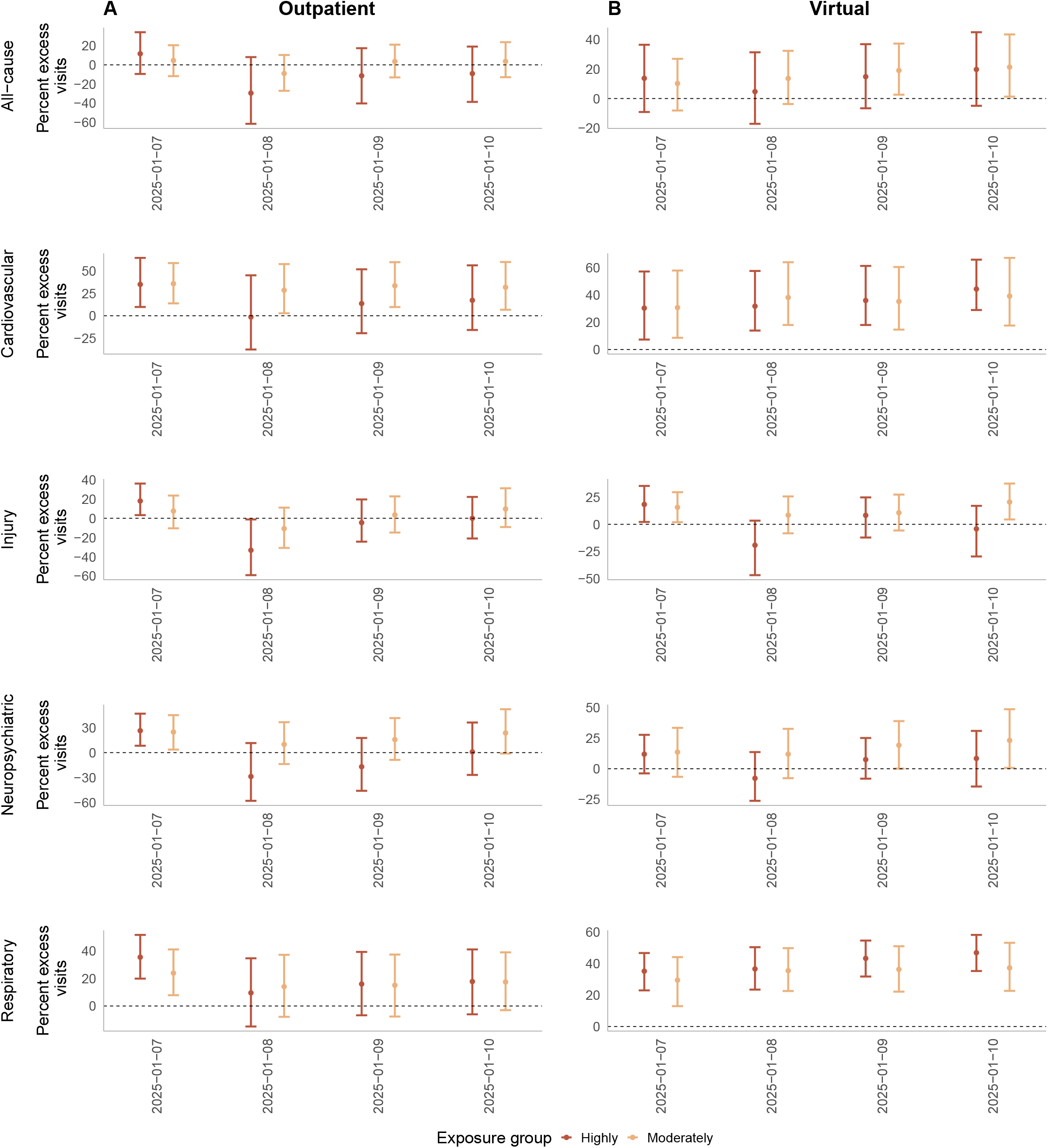
Estimated change in the percent of (A) outpatient and (B) virtual acute care visits for all-cause, cardiovascular, injury, neuropsychiatric, and respiratory endpoints at Kaiser Permanente Southern California (KPSC) in the four days following the January 7, 2025 ignition of the LA Fires. We used the maximum wildfire burn zone reached by an LA or Ventura County wildfire as of January 16, 2025 to define exposure. Highly exposed members resided in a census tract located <20km burn zones, and moderately exposed members lived in tracts >20km but within LA County. Results from an interrupted time series model using KPSC electronic health record data from November to January 2022–2025, with daily maximum and minimum temperature and humidity, wind velocity, and surface downward shortwave radiation and weekly wastewater surveillance data on levels of three respiratory viruses as covariates. We employed a Monte Carlo simulation approach, performing 1,000 model iterations to estimate 95% empirical confidence intervals (eCIs) for the predictions.

On January 7, we observed excess outpatient and virtual cardiovascular and respiratory visits in highly- and moderately-exposed groups. Virtual cardiovascular and respiratory visits remained elevated through January 10, were unchanged on January 11-12 (weekend), then rose on January 13, in both highly- and moderately-exposed groups. Outpatient cardiovascular visits in the moderately-exposed group followed a similar trend, but outpatient respiratory visits did not.

On January 7, outpatient injury visits were 18% (n=53, 95% eCI: 10, 107) above expected in the highly-exposed group, and injury virtual visits were 18% and 16% above expected in the highly- and moderately-exposed groups, respectively (**eFigure 2, eTables 1-8**). Outpatient neuropsychiatric visits were approximately 25% higher than expected on January 7, totaling 279 (95% eCI: 88, 498) and 1124 (95% eCI: 165, 2044) excess visits in the highly- and moderately-exposed groups, respectively.

When extending follow-up to two weeks post-January 7, outpatient cardiovascular, neuropsychiatric, and respiratory visits exceeded expected levels by 21-45% from January 13-17 (**eTables 5-6**) in both highly- and moderately-exposed groups. Cardiovascular and respiratory virtual visits also remained 25-41% above expected levels through January 17 in both highly- and moderately-exposed groups (**eTables 7-8**).

Sensitivity analysis indicated that respiratory-related virtual visits in the minimally-exposed group were 29% (95% eCI: 13, 45) higher than expected during the week following January 7, compared to 34% for moderately exposed and 41% for the highly exposed groups (**eFigure 3**).

We estimated the impacts of the LA Fire using a simplified difference-in-differences framework that did not account for complex seasonal trends or time-varying covariates. These analyses found contradictory results, highlighting the utility of employing flexible ITS methods for rapid health impact assessments (**eTable 9**).

## Discussion

The LA Fires ignited on January 7, 2025, and evacuations, power outages, displacement, and devastating loss of property and lives ensued.^1^ In the week following ignition, we observed approximately 35% increases in virtual care seeking for cardiovascular- and respiratory-related conditions among highly- or moderately-exposed KPSC members. Short-term injury and neuropsychiatric care-seeking also increased.

Our findings align with prior work finding increased acute emergency and inpatient cardiorespiratory and mental health care use following wildfire smoke exposure.^8-10^ Studies of wildfire disasters have also documented increased injuries, including from motor vehicle crashes during the Thomas Fire evacuation.^11^

Virtual versus in-person care-seeking increased substantially more immediately following the LA Fires, suggesting healthcare systems should prioritize adequate virtual care staffing during climate events.^12^ We also identified potential catch-up outpatient care for cardiorespiratory and neuropsychiatric concerns from January 12-16 after the worst of the smoke had cleared.

Our study had limitations. We did not differentiate between smoke and burn zone exposure. Both exposures impact members of persistently disadvantaged groups differentially.^13-15^ Early reports indicate many killed by the LA fires were older adults and disabled people.^16^ Our time-series analyses did not assess individual or community-level vulnerability factors, like housing quality or who successfully evacuated. Because it takes KPSC longer to reconcile cause-specific emergency department and inpatient visit data accurately, we restricted our initial study to outpatient and virtual visits.

Climate hazards will increasingly impact US cities. Here, we demonstrated that EHR data can rapidly inform healthcare delivery needs and population health impacts after such events. We identified substantial increases in acute care visits, driven primarily by virtual care-seeking, the role of which will likely grow in serving patients during disruptive climate events.

## Data Availability

Anonymized data that support the findings of this study may be made available from the investigative team in the following conditions: (1) agreement to collaborate with the study team on all publications, (2) provision of external funding for administrative and investigator time necessary for this collaboration, (3) demonstration that the external investigative team is qualified and has documented evidence of training for human subjects protections, and (4) agreement to abide by the terms outlined in data use agreements between institutions.

## Acknowledgments

T.B., J.A.C., T.B.F, and S.Y.T. were supported by the NIH National Institute on Aging (NIA) grant R01AG071024. J.A.C. was also supported by the NIH National Institute for Environmental Health Sciences (NIEHS) grant P30ES007033. T.B. was also supported by the NIA (RF1AG080948) and NIEHS P20MD019994. Neither NIA nor NIEHS had any role in the design and conduct of the study; collection, management, analysis, and interpretation of the data; preparation, review, or approval of the manuscript; and decision to submit the manuscript for publication. The authors thank Ms. Elizabeth Blake for her assistance in preparing tables and figures.

