## Supplemental Materials for "The 2025 Los Angeles Wildfires and Outpatient Acute Healthcare Utilization"

**The 2025 Los Angeles Wildfires and Short-Term Outpatient Acute Healthcare Utilization Supplemental Materials**

### eMethods for the 2-stage interrupted time-series design.

This study uses an observational two-stage interrupted time series design using machine learning methods to estimate the number of visits attributable to wildfire exposure. The first stage generates counterfactual trends of the expected number of visits for each of the exposure groups and type of encounter in the absence of the wildfires using a hybrid Prophet-Extreme Gradient Boosting (XGBoost) model. We use the pre-wildfire period, November-January periods from November 1, 2022 to January 6, 2025, for training and testing of the modelling framework and apply the model to the post-wildfire ignition period (January 7, 2025 to January 20, 2025). In the second stage, we used the modelled estimates of the counterfactual trend to estimate excess visits from each level of wildfire exposure (minimally, moderately, and highly).

**Background on modeling framework**

This machine learning approach combines the Prophet and XGBoost algorithms into a robust forecasting framework. First, Prophet decomposes the time series into trend, seasonality, and holiday components, which provides a structured representation of temporal patterns. Then, XGBoost augments this framework by modeling the residuals from the Prophet model through an ensemble of decision trees, minimizing the regularized objective function that balances prediction error and model complexity. The hybrid implementation leverages Prophet's strength in capturing explicit temporal patterns while using XGBoost's ability to identify complex relationships in the residuals and additional features.

Prophet is an additive regression model developed by Facebook for forecasting time series data. It decomposes time series into three main components: trend, seasonality, and holidays. The trend component is modeled using a logistic growth curve to accommodate saturated growth patterns. The holiday component allows for the incorporation of specific events or occasions that can influence the time series data.[1] The model can be expressed as:

$$y(t) = g(t) + s(t) + h(t) + \varepsilon_t$$

Where $g(t)$ represents the trend function, $s(t)$ is the seasonal component that captures periodic changes, $h(t)$ accounts for the effects of holidays, and $\varepsilon_t$ is the error term.

XGBoost is an advanced implementation of gradient boosting, where predictive models are built as an ensemble of weak learners, typically decision trees. It operates by consecutively adding predictors to an ensemble, with each new predictor correcting its predecessor's errors through gradient descent optimization. XGBoost minimizes a regularized objective function that balances prediction error and model complexity, ensuring both accuracy and simplicity in the final model. The predictive model in XGBoost is formalized as follows:

$$\hat{y}_i = \sum_{k=1}^{K} f_k(x_i), \quad f_k \in \Phi$$

where $\hat{y}_i$ is the prediction for the $i$-th instance, $x_i$ represents the features of the $i$-th instance, $K$ is the number of trees in the model, $f_k$ represents the individual decision trees, and $\Phi$ is the space of all possible regression trees.

**Statistical modeling in this analysis**

As we are using data from partial years (November-January) with strong weekend trends, we included indicators for U.S. federal holidays, time period (Nov 2022-Jan 2023, Nov 2023-Jan 2024, Nov 2024-Jan 2025), weekday/weekend, day of the week, month, and year in our model. We specifically removed certain holidays (Inauguration Day, Pulaski's Birthday, Juneteenth, and Decoration Memorial Day) and added an indicator for business closed holidays at Kaiser Permanente Southern California (Christmas Day, New Year's Day, Martin Luther King Day, Independence Day). We included covariates for daily maximum and minimum temperature and humidity, wind velocity, and surface downward shortwave radiation from gridMET[2] as well as weekly wastewater surveillance data on levels of three respiratory viruses: flu, respiratory syncytial virus (RSV), and SARS-CoV-2 [3] to improve model predictions. Due to differing seasonal patterns for respiratory Virtual visits in Fall 2022 from the rest of the time series, models for these visits were restricted to January 2023 and later.

For our analysis, we used a time series split approach dividing the dataset into training and testing periods. Specifically, we held out ~30% of pre-wildfire days for testing while using the remaining data for training. Within the training data, we implemented a time series cross-validation framework with 8 resampling slices using an assessment period ~20% of the training dataset for each fold. This approach maintains the temporal dependencies in the data while ensuring robust model validation.

We employed Prophet with XGBoost (Prophet Boost) using tunable parameters. For the XGBoost component, we tuned key parameters including number of variables per split (mtry: 2-10), minimum node size (min_n: 15-30), tree depth (3-8), learning rate (0.001-0.1), loss reduction ($10^{-5}$ to 10), and early stopping iterations (10-50) for the XGBoost component. The tuning parameters for the XGBoost component were further specified within these ranges to find the best fit for some of the encounter types and causes. The tuning process used a space-filling design with 100 combinations.

We identified the optimal parameter set by minimizing the root mean square error (RMSE) across the cross-validation folds. We then used these optimal parameters to fit the final model on the entire training dataset and evaluated its performance on both the training data and the held-out test data using multiple metrics including mean absolute error (MAE), RMSE, mean absolute percentage error (MAPE), and R-squared.

We then used the optimal parameters to predict the number of visits for the whole study period. We employed a Monte Carlo simulation approach performing 1,000 model iterations to estimate 95% empirical confidence intervals (eCIs) for the predictions.[4] The eCIs were derived from the empirical distribution of the estimated number of visits for each day, taking the 2.5$^{th}$ and 97.5$^{th}$ percentiles as the lower and upper bounds.

To estimate the effect of the LA Fires for each encounter type, cause and wildfire exposure category, we calculated the difference between the observed and expected number of visits based on the model predictions. The number of excess visits was estimated for each day in the post-wildfire period for each encounter type, cause and exposure category. We estimated eCIs for excess visits by taking the difference between observed and the upper and lower bound of the prediction, which came from our Monte Carlo simulation. The total number of Kaiser beneficiaries in each exposure group was used to calculate the excess number of visits per 1,000 beneficiaries. A total number of visits was estimated by taking a sum of the excess visits and its upper and lower bounds across the week following the initial wildfires.

All statistical analyses were conducted with R software (version 4.4.1) using the *tidymodels tidyverse, modeltime, timetk, tictoc, metrics*, *fst* and *data.table* packages and Python version 3.12.2. The datasets and R scripts used in this study are publicly available on GitHub.

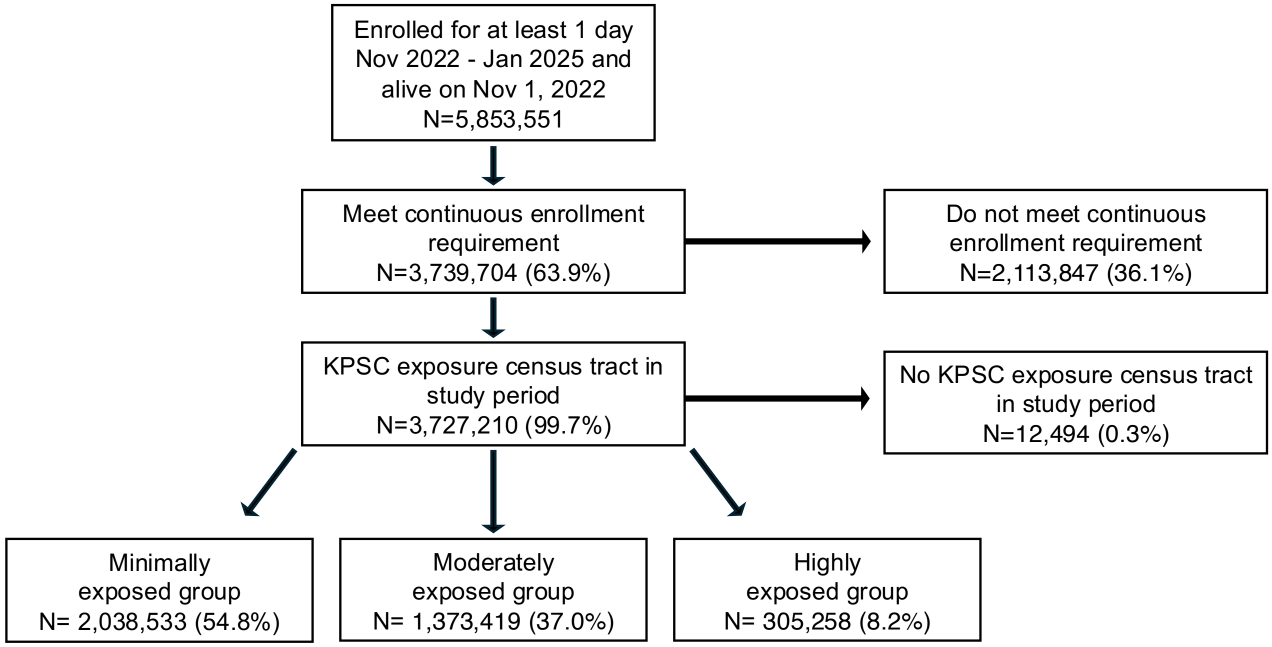


**eFigure 1. Flow diagram depicting exclusion criteria for members of the KPSC study population, 2022-2025.** To be included, KPSC members had to be enrolled from November to January 2022–2025, allowing 30-day enrollment gaps. The KPSC catchment counties included Imperial, Kern, Los Angeles, Orange, Riverside, San Bernardino, San Diego, San Luis Obispo, Santa Barbara, and Ventura. We excluded KPSC members who did not have a residential address in the KPSC catchment at baseline.
KPSC, Kaiser Permanente Southern California

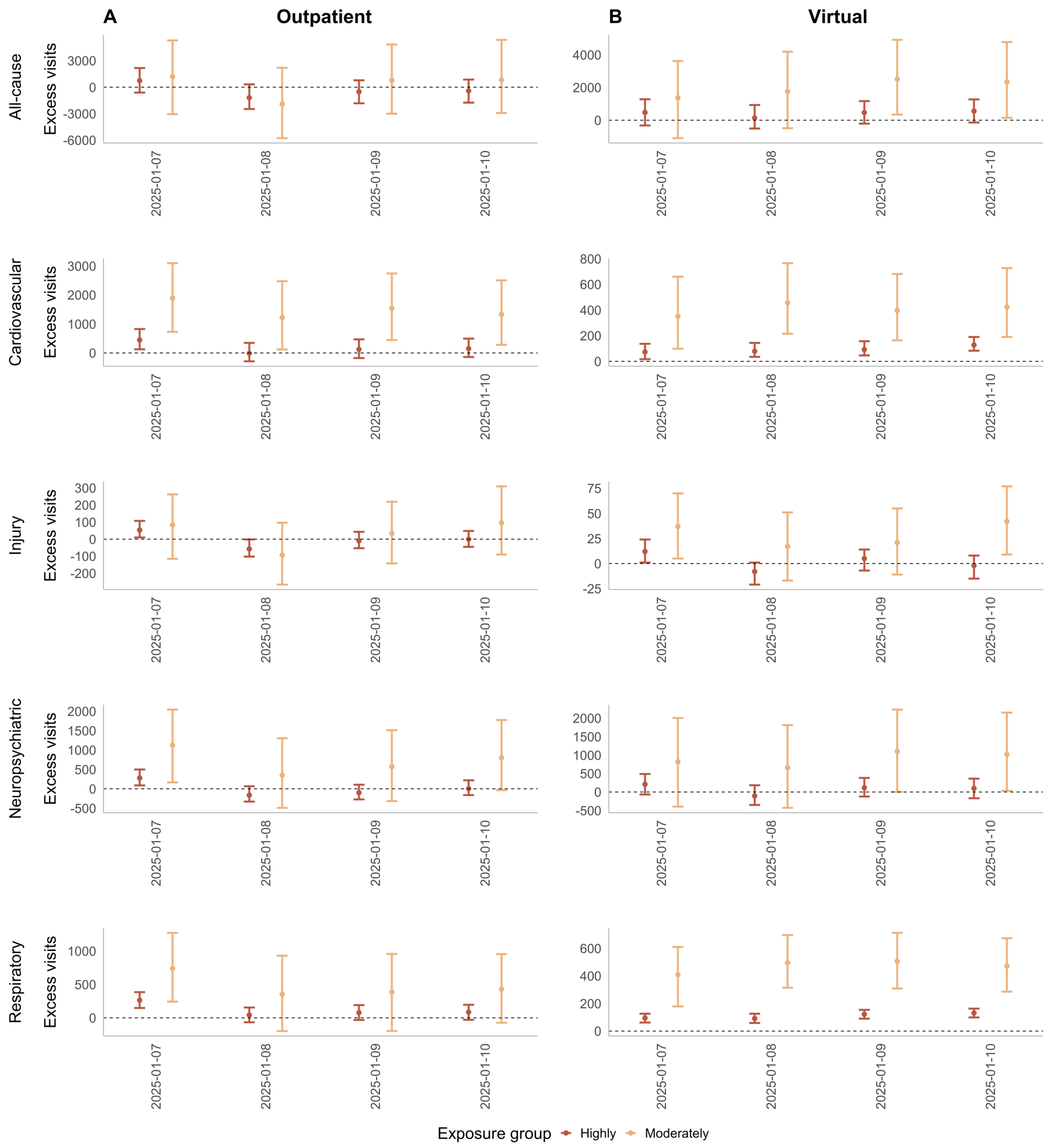


**eFigure 2. Estimated change in the count of (A) outpatient and (B) virtual acute care visits for all-cause, cardiovascular, injury, neuropsychiatric, and respiratory endpoints at Kaiser Permanente Southern California (KPSC) in the four days following the January 7, 2025 ignition of the LA Fires.** We used the maximum wildfire burn zone reached by an LA or Ventura County wildfire as of January 16, 2025 to define exposure. Highly exposed members resided in a census tract located <20km burn zones, and moderately exposed members lived in tracts ≥20km but within LA County. Results from an interrupted time series model using KPSC electronic health record data from November to January 2022–2025, with daily maximum and minimum temperature and humidity, wind velocity, and surface downward shortwave radiation and weekly wastewater surveillance data on levels of three respiratory viruses as covariates. We employed a Monte Carlo simulation approach, performing 1,000 model iterations to estimate 95% empirical confidence intervals (eCIs) for the predictions.

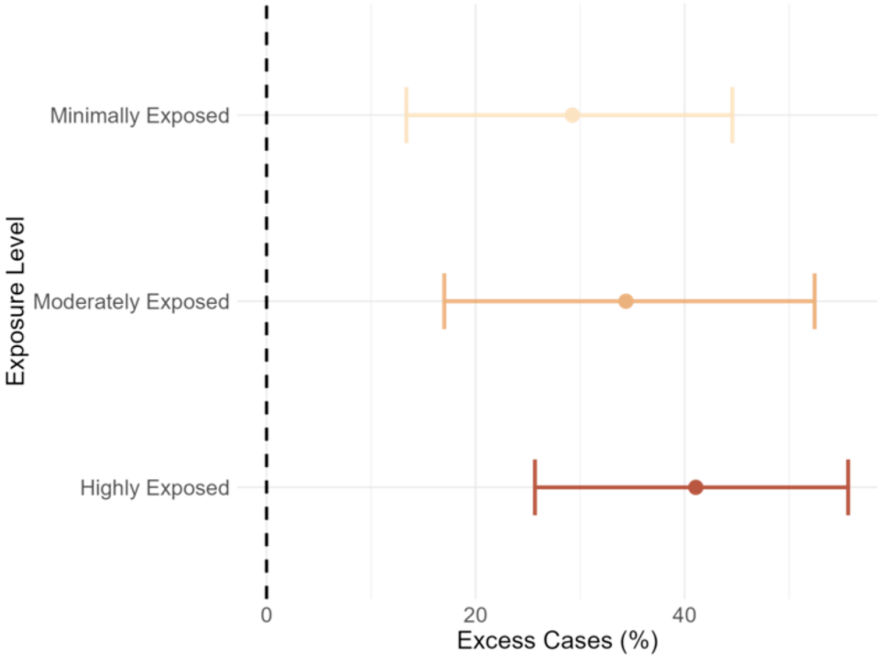


**eFigure 3. Estimated change in the percent of respiratory-related virtual care visits at Kaiser Permanente Southern California (KPSC) in the week following the January 7, 2025 ignition of the 2025 LA Fires.** We used the maximum wildfire burn zone reached by an LA or Ventura County wildfire as of January 16, 2025 to define exposure. Highly exposed members resided in a census tract located <20km burn zones, moderately exposed members lived in tracts ≥20km but within LA County, and minimally exposed members lived in tracts ≥20km in other KPSC catchment counties (Imperial, Kern, Orange, Riverside, San Bernardino, San Diego, San Luis Obispo, Santa Barbara, and Ventura). Results from an interrupted time series model using KPSC electronic health record data from November to January 2022–2025, with daily maximum and minimum temperature and humidity, wind velocity, and surface downward shortwave radiation and weekly wastewater surveillance data on levels of three respiratory viruses as covariates. We employed a Monte Carlo simulation approach, performing 1,000 model iterations to estimate 95% empirical confidence intervals (eCIs) for the predictions.

**eTable 1. Estimated change in the count of outpatient acute care visits for all-cause, cardiovascular, injury, neuropsychiatric, and respiratory endpoints among Kaiser Permanente Southern California (KPSC) highly-exposed members (n=305,258) in the two weeks following the January 7, 2025 ignition of the LA Fires.**

| | | *n (95% eCI)* | | | | |
|---|---|---|---|---|---|---|
| **Date** | **Weekday** | **All-cause** | **Cardiovascular** | **Injury** | **Neuropsychiatric** | **Respiratory** |
| 2025-01-07 | Tuesday | 741 (-607, 2170) | 447 (126, 824) | 53 (10, 107) | 279 (88, 498) | 264 (148, 385) |
| 2025-01-08 | Wednesday | -1182 (-2465, 323) | -11 (-289, 347) | -57 (-102, -2) | -164 (-331, 65) | 43 (-66, 155) |
| 2025-01-09 | Thursday | -517 (-1820, 784) | 124 (-176, 470) | -10 (-53, 43) | -101 (-274, 105) | 78 (-32, 191) |
| 2025-01-10 | Friday | -409 (-1746, 864) | 152 (-140, 495) | 0 (-45, 48) | 5 (-163, 219) | 85 (-29, 196) |
| 2025-01-11 | Saturday | 273 (-950, 1628) | -141 (-386, 176) | -3 (-43, 51) | -75 (-240, 143) | -46 (-149, 68) |
| 2025-01-12 | Sunday | 458 (-746, 1916) | -154 (-395, 180) | -10 (-50, 48) | -56 (-226, 157) | -38 (-142, 81) |
| 2025-01-13 | Monday | 1174 (-97, 2642) | 464 (200, 818) | 30 (-12, 86) | 221 (51, 442) | 333 (225, 456) |
| 2025-01-14 | Tuesday | 701 (-583, 2148) | 412 (141, 761) | 24 (-16, 80) | 206 (40, 419) | 232 (123, 359) |
| 2025-01-15 | Wednesday | 923 (-333, 2414) | 381 (122, 728) | 15 (-29, 69) | 275 (108, 493) | 212 (107, 333) |
| 2025-01-16 | Thursday | 1317 (-159, 3038) | 326 (61, 692) | 57 (14, 119) | 227 (53, 464) | 246 (137, 371) |
| 2025-01-17 | Friday | 923 (-432, 2336) | 214 (-61, 555) | 52 (12, 98) | 171 (-4, 379) | 154 (43, 274) |
| 2025-01-18 | Saturday | 443 (-800, 1744) | -143 (-391, 187) | 5 (-34, 48) | -61 (-236, 146) | -54 (-159, 66) |
| 2025-01-19 | Sunday | 578 (-641, 1879) | -141 (-390, 182) | -8 (-47, 38) | -71 (-242, 127) | -44 (-150, 74) |
| 2025-01-20 | Monday | -3951 (-5388, -2234) | -708 (-964, -360) | -195 (-237, -134) | -384 (-557, -148) | -385 (-494, -266) |

We used the maximum wildfire burn zone reached by an LA or Ventura County wildfire as of January 16, 2025 to define exposure. Highly exposed members resided in a census tract located <20km burn zones. Results from an interrupted time series model using KPSC electronic health record data from November to January 2022–2025, with daily maximum and minimum temperature and humidity, wind velocity, and surface downward shortwave radiation and weekly wastewater surveillance data on levels of three respiratory viruses as covariates. We employed a Monte Carlo simulation approach, performing 1,000 model iterations to estimate 95% empirical confidence intervals (eCIs) for the predictions.

**eTable 2. Estimated change in the count of outpatient acute care visits for all-cause, cardiovascular, injury, neuropsychiatric, and respiratory endpoints among Kaiser Permanente Southern California (KPSC) moderately-exposed members (n=1,373,419) in the two weeks following the January 7, 2025 ignition of the LA Fires.**

| *n (95% eCI)* | | | | | | |
|---|---|---|---|---|---|---|
| **Date** | **Weekday** | **All-cause** | **Cardiovascular** | **Injury** | **Neuropsychiatric** | **Respiratory** |
| 2025-01-07 | Tuesday | 1200 (-3047, 5298) | 1890 (729, 3098) | 83 (-115, 262) | 1124 (165, 2044) | 739 (243, 1272) |
| 2025-01-08 | Wednesday | -1913 (-5760, 2201) | 1223 (120, 2475) | -93 (-266, 96) | 352 (-492, 1306) | 352 (-198, 931) |
| 2025-01-09 | Thursday | 795 (-2991, 4835) | 1539 (445, 2743) | 34 (-142, 219) | 572 (-318, 1514) | 386 (-196, 957) |
| 2025-01-10 | Friday | 827 (-2920, 5358) | 1329 (281, 2504) | 96 (-90, 309) | 804 (-36, 1775) | 428 (-73, 954) |
| 2025-01-11 | Saturday | 442 (-2946, 4795) | -399 (-1329, 701) | -21 (-186, 182) | -176 (-995, 762) | -221 (-646, 250) |
| 2025-01-12 | Sunday | 546 (-3376, 5333) | -436 (-1391, 789) | -34 (-202, 190) | -191 (-970, 815) | -252 (-712, 253) |
| 2025-01-13 | Monday | 5278 (1332, 9791) | 1931 (914, 3146) | 241 (59, 462) | 1289 (484, 2273) | 1037 (510, 1536) |
| 2025-01-14 | Tuesday | 2223 (-1511, 7092) | 1802 (878, 3017) | 118 (-66, 338) | 1307 (467, 2307) | 709 (220, 1256) |
| 2025-01-15 | Wednesday | 2686 (-1184, 7155) | 1402 (368, 2611) | 135 (-69, 357) | 1179 (375, 2105) | 694 (192, 1218) |
| 2025-01-16 | Thursday | 5839 (1876, 11027) | 1679 (644, 2830) | 266 (79, 481) | 1225 (390, 2207) | 817 (361, 1340) |
| 2025-01-17 | Friday | 3066 (-642, 7599) | 1047 (-13, 2169) | 110 (-60, 303) | 990 (179, 1862) | 521 (104, 986) |
| 2025-01-18 | Saturday | 1071 (-2557, 5536) | -452 (-1389, 689) | -29 (-198, 155) | -202 (-1010, 770) | -174 (-565, 331) |
| 2025-01-19 | Sunday | 1382 (-2264, 5967) | -479 (-1470, 629) | -30 (-198, 160) | -257 (-1100, 710) | -163 (-572, 337) |
| 2025-01-20 | Monday | -18042 (-21902, -13060) | -2960 (-3954, -1820) | -796 (-975, -593) | -1774 (-2567, -893) | -1808 (-2262, -1279) |

We used the maximum wildfire burn zone reached by an LA or Ventura County wildfire as of January 16, 2025 to define exposure. Moderately exposed members lived in tracts ≥20km from a burn zone but within LA County. Results from an interrupted time series model using KPSC electronic health record data from November to January 2022–2025, with daily maximum and minimum temperature and humidity, wind velocity, and surface downward shortwave radiation and weekly wastewater surveillance data on levels of three respiratory viruses as covariates. We employed a Monte Carlo simulation approach, performing 1,000 model iterations to estimate 95% empirical confidence intervals (eCIs) for the predictions.

**eTable 3. Estimated change in the count of virtual acute care visits for all-cause, cardiovascular, injury, neuropsychiatric, and respiratory endpoints among Kaiser Permanente Southern California (KPSC) highly-exposed members (n=305,258) in the two weeks following the January 7, 2025 ignition of the LA Fires.**

| | | *n (95% eCI)* | | | | |
|---|---|---|---|---|---|---|
| **Date** | **Weekday** | **All-cause** | **Cardiovascular** | **Injury** | **Neuropsychiatric** | **Respiratory** |
| 2025-01-07 | Tuesday | 482 (-321, 1282) | 73 (17, 137) | 12 (1, 24) | 209 (-67, 488) | 95 (62, 126) |
| 2025-01-08 | Wednesday | 140 (-507, 931) | 79 (35, 144) | -8 (-21, 1) | -104 (-351, 183) | 91 (59, 126) |
| 2025-01-09 | Thursday | 471 (-209, 1171) | 92 (46, 157) | 5 (-7, 14) | 115 (-123, 382) | 122 (90, 154) |
| 2025-01-10 | Friday | 560 (-140, 1275) | 128 (83, 190) | -2 (-15, 8) | 99 (-170, 362) | 131 (99, 163) |
| 2025-01-11 | Saturday | 317 (-323, 1005) | -11 (-55, 46) | 1 (-10, 11) | 10 (-235, 279) | 33 (0, 65) |
| 2025-01-12 | Sunday | 307 (-331, 1049) | -10 (-53, 53) | -6 (-16, 3) | 25 (-226, 307) | 32 (1, 60) |
| 2025-01-13 | Monday | 804 (117, 1623) | 78 (35, 143) | 15 (3, 25) | 281 (30, 544) | 102 (70, 129) |
| 2025-01-14 | Tuesday | 586 (-101, 1385) | 73 (29, 137) | 7 (-4, 18) | 217 (-36, 503) | 61 (29, 89) |
| 2025-01-15 | Wednesday | 524 (-164, 1300) | 76 (31, 140) | 7 (-5, 17) | 198 (-59, 469) | 104 (72, 132) |
| 2025-01-16 | Thursday | 903 (191, 1840) | 110 (67, 181) | 3 (-7, 15) | 353 (92, 685) | 92 (59, 120) |
| 2025-01-17 | Friday | 507 (-172, 1177) | 51 (9, 113) | -11 (-22, -1) | 193 (-38, 452) | 65 (35, 93) |
| 2025-01-18 | Saturday | 263 (-402, 937) | -18 (-61, 42) | -2 (-12, 8) | 61 (-178, 315) | 10 (-19, 38) |
| 2025-01-19 | Sunday | 310 (-343, 977) | -11 (-52, 49) | 4 (-7, 13) | 27 (-214, 282) | 22 (-8, 53) |
| 2025-01-20 | Monday | -1273 (-1990, -376) | -138 (-182, -69) | -41 (-52, -29) | -327 (-585, 5) | -89 (-121, -56) |

We used the maximum wildfire burn zone reached by an LA or Ventura County wildfire as of January 16, 2025 to define exposure. Highly exposed members resided in a census tract located <20km burn zones. Results from an interrupted time series model using KPSC electronic health record data from November to January 2022–2025, with daily maximum and minimum temperature and humidity, wind velocity, and surface downward shortwave radiation and weekly wastewater surveillance data on levels of three respiratory viruses as covariates. We employed a Monte Carlo simulation approach, performing 1,000 model iterations to estimate 95% empirical confidence intervals (eCIs) for the predictions.

**eTable 4. Estimated change in the count of virtual acute care visits for all-cause, cardiovascular, injury, neuropsychiatric, and respiratory endpoints among Kaiser Permanente Southern California (KPSC) moderately-exposed members (n=1,373,419) in the two weeks following the January 7, 2025 ignition of the LA Fires.**

| | | *n (95% eCI)* | | | | |
|---|---|---|---|---|---|---|
| **Date** | **Weekday** | **All-cause** | **Cardiovascular** | **Injury** | **Neuropsychiatric** | **Respiratory** |
| 2025-01-07 | Tuesday | 1368 (-1093, 3625) | 351 (98, 660) | 37 (5, 70) | 818 (-396, 2005) | 408 (179, 610) |
| 2025-01-08 | Wednesday | 1760 (-486, 4194) | 457 (215, 766) | 17 (-17, 51) | 659 (-429, 1812) | 495 (315, 696) |
| 2025-01-09 | Thursday | 2512 (347, 4915) | 397 (164, 681) | 21 (-11, 55) | 1101 (4, 2232) | 506 (309, 712) |
| 2025-01-10 | Friday | 2345 (147, 4780) | 424 (190, 727) | 42 (9, 77) | 1022 (32, 2151) | 471 (286, 673) |
| 2025-01-11 | Saturday | 1518 (-762, 3957) | 3 (-204, 265) | 25 (-7, 59) | 313 (-659, 1371) | 119 (-67, 318) |
| 2025-01-12 | Sunday | 1515 (-636, 4248) | -10 (-223, 303) | 16 (-17, 54) | 252 (-743, 1408) | 90 (-78, 268) |
| 2025-01-13 | Monday | 3700 (1358, 6414) | 373 (148, 684) | 56 (22, 95) | 1367 (218, 2665) | 525 (348, 710) |
| 2025-01-14 | Tuesday | 1903 (-374, 4740) | 379 (160, 688) | 12 (-22, 51) | 837 (-199, 2069) | 373 (194, 557) |
| 2025-01-15 | Wednesday | 2212 (-256, 4808) | 300 (80, 611) | 20 (-13, 57) | 815 (-315, 2060) | 348 (163, 541) |
| 2025-01-16 | Thursday | 3389 (1120, 6411) | 367 (139, 676) | 19 (-13, 63) | 1262 (170, 2566) | 489 (323, 656) |
| 2025-01-17 | Friday | 2068 (-32, 4480) | 219 (7, 499) | 27 (-7, 62) | 774 (-332, 1944) | 318 (135, 480) |
| 2025-01-18 | Saturday | 1268 (-908, 3627) | -22 (-221, 269) | 5 (-28, 40) | 257 (-836, 1470) | 70 (-90, 224) |
| 2025-01-19 | Sunday | 1163 (-1043, 3501) | -25 (-228, 256) | 13 (-19, 48) | 205 (-912, 1382) | 120 (-50, 293) |
| 2025-01-20 | Monday | -5488 (-7724, -2650) | -611 (-839, -310) | -113 (-147, -71) | -1353 (-2522, -238) | -286 (-474, -93) |

We used the maximum wildfire burn zone reached by an LA or Ventura County wildfire as of January 16, 2025 to define exposure. Moderately exposed members lived in tracts ≥20km from a burn zone but within LA County. Results from an interrupted time series model using KPSC electronic health record data from November to January 2022–2025, with daily maximum and minimum temperature and humidity, wind velocity, and surface downward shortwave radiation and weekly wastewater surveillance data on levels of three respiratory viruses as covariates. We employed a Monte Carlo simulation approach, performing 1,000 model iterations to estimate 95% empirical confidence intervals (eCIs) for the predictions.

**eTable 5. Estimated change in the percent of outpatient acute care visits for all-cause, cardiovascular, injury, neuropsychiatric, and respiratory endpoints among Kaiser Permanente Southern California (KPSC) highly-exposed members (n=305,258) in the two weeks following the January 7, 2025 ignition of the LA Fires.**

| *Percent (95% eCI)* | | | | | | |
|---|---|---|---|---|---|---|
| **Date** | **Weekday** | **All-cause** | **Cardiovascular** | **Injury** | **Neuropsychiatric** | **Respiratory** |
| 2025-01-07 | Tuesday | 12% (-10%, 34%) | 35% (10%, 64%) | 18% (3%, 36%) | 26% (8%, 47%) | 35% (20%, 51%) |
| 2025-01-08 | Wednesday | -30% (-62%, 8%) | -1% (-37%, 45%) | -33% (-59%, -1%) | -29% (-58%, 11%) | 9% (-15%, 35%) |
| 2025-01-09 | Thursday | -11% (-40%, 17%) | 14% (-19%, 52%) | -5% (-24%, 20%) | -17% (-46%, 18%) | 16% (-7%, 39%) |
| 2025-01-10 | Friday | -9% (-39%, 19%) | 17% (-16%, 56%) | 0% (-21%, 22%) | 1% (-27%, 36%) | 18% (-6%, 41%) |
| 2025-01-11 | Saturday | 41% (-142%, 244%) | -198% (-543%, 249%) | -9% (-150%, 177%) | -116% (-369%, 219%) | -178% (-574%, 262%) |
| 2025-01-12 | Sunday | 477% (-777%, 1995%) | -1181% (-3040%, 1387%) | -317% (-1651%, 1612%) | -295% (-1191%, 827%) | -225% (-833%, 478%) |
| 2025-01-13 | Monday | 20% (-2%, 45%) | 38% (16%, 67%) | 12% (-5%, 36%) | 25% (6%, 50%) | 45% (31%, 62%) |
| 2025-01-14 | Tuesday | 11% (-9%, 34%) | 33% (11%, 61%) | 9% (-6%, 30%) | 21% (4%, 42%) | 32% (17%, 50%) |
| 2025-01-15 | Wednesday | 15% (-5%, 39%) | 33% (11%, 64%) | 6% (-12%, 28%) | 27% (11%, 48%) | 35% (17%, 54%) |
| 2025-01-16 | Thursday | 22% (-3%, 52%) | 30% (6%, 63%) | 21% (5%, 43%) | 25% (6%, 51%) | 38% (21%, 57%) |
| 2025-01-17 | Friday | 17% (-8%, 43%) | 23% (-7%, 60%) | 20% (5%, 38%) | 23% (-1%, 50%) | 29% (8%, 51%) |
| 2025-01-18 | Saturday | 71% (-128%, 279%) | -281% (-766%, 366%) | 16% (-102%, 147%) | -78% (-302%, 187%) | -360% (-1059%, 440%) |
| 2025-01-19 | Sunday | 622% (-689%, 2021%) | -1760% (-4875%, 2280%) | -382% (-2345%, 1910%) | -886% (-3029%, 1591%) | -403% (-1366%, 674%) |
| 2025-01-20 | Monday | -764% (-1042%, -432%) | -3078% (-4192%, -1565%) | -1771% (-2153%, -1219%) | -147% (-213%, -57%) | -3207% (-4116%, -2216%) |

We used the maximum wildfire burn zone reached by an LA or Ventura County wildfire as of January 16, 2025 to define exposure. Highly exposed members resided in a census tract located <20km burn zones. Results from an interrupted time series model using KPSC electronic health record data from November to January 2022–2025, with daily maximum and minimum temperature and humidity, wind velocity, and surface downward shortwave radiation and weekly wastewater surveillance data on levels of three respiratory viruses as covariates. We employed a Monte Carlo simulation approach, performing 1,000 model iterations to estimate 95% empirical confidence intervals (eCIs) for the predictions.

**eTable 6. Estimated change in the percent of outpatient acute care visits for all-cause, cardiovascular, injury, neuropsychiatric, and respiratory endpoints among Kaiser Permanente Southern California (KPSC) moderately-exposed members (n=1,373,419) in the two weeks following the January 7, 2025 ignition of the LA Fires.**

| | | *Percent (95% eCI)* | | | | |
|---|---|---|---|---|---|---|
| **Date** | **Weekday** | **All-cause** | **Cardiovascular** | **Injury** | **Neuropsychiatric** | **Respiratory** |
| 2025-01-07 | Tuesday | 5% (-12%, 20%) | 36% (14%, 59%) | 7% (-10%, 24%) | 25% (4%, 45%) | 24% (8%, 41%) |
| 2025-01-08 | Wednesday | -9% (-27%, 10%) | 28% (3%, 58%) | -11% (-31%, 11%) | 10% (-14%, 36%) | 14% (-8%, 37%) |
| 2025-01-09 | Thursday | 3% (-13%, 21%) | 33% (10%, 60%) | 4% (-15%, 23%) | 16% (-9%, 41%) | 15% (-8%, 37%) |
| 2025-01-10 | Friday | 4% (-13%, 24%) | 32% (7%, 60%) | 10% (-9%, 31%) | 24% (-1%, 52%) | 17% (-3%, 39%) |
| 2025-01-11 | Saturday | 18% (-117%, 191%) | -182% (-607%, 320%) | -33% (-287%, 280%) | -43% (-244%, 187%) | -131% (-382%, 148%) |
| 2025-01-12 | Sunday | 125% (-776%, 1226%) | -1505% (-4797%, 2719%) | -215% (-1264%, 1188%) | -239% (-1212%, 1018%) | -360% (-1016%, 361%) |
| 2025-01-13 | Monday | 20% (5%, 37%) | 39% (18%, 63%) | 22% (5%, 42%) | 30% (11%, 54%) | 34% (17%, 50%) |
| 2025-01-14 | Tuesday | 8% (-6%, 27%) | 35% (17%, 59%) | 10% (-6%, 30%) | 28% (10%, 49%) | 23% (7%, 41%) |
| 2025-01-15 | Wednesday | 10% (-5%, 28%) | 31% (8%, 58%) | 12% (-6%, 33%) | 27% (8%, 47%) | 25% (7%, 45%) |
| 2025-01-16 | Thursday | 23% (7%, 43%) | 36% (14%, 61%) | 23% (7%, 42%) | 29% (9%, 52%) | 29% (13%, 48%) |
| 2025-01-17 | Friday | 13% (-3%, 32%) | 27% (0%, 55%) | 11% (-6%, 31%) | 27% (5%, 51%) | 21% (4%, 40%) |
| 2025-01-18 | Saturday | 45% (-107%, 231%) | -319% (-978%, 485%) | -53% (-359%, 282%) | -49% (-246%, 188%) | -146% (-475%, 278%) |
| 2025-01-19 | Sunday | 337% (-552%, 1455%) | -1228% (-3768%, 1613%) | -248% (-1651%, 1330%) | -352% (-1507%, 973%) | -286% (-1003%, 592%) |
| 2025-01-20 | Monday | -766% (-930%, -554%) | -5193% (-6938%, -3194%) | -1502% (-1839%, -1119%) | -151% (-218%, -76%) | -3410% (-4268%, -2412%) |

We used the maximum wildfire burn zone reached by an LA or Ventura County wildfire as of January 16, 2025 to define exposure. Moderately exposed members lived in tracts ≥20km from a burn zone but within LA County. Results from an interrupted time series model using KPSC electronic health record data from November to January 2022–2025, with daily maximum and minimum temperature and humidity, wind velocity, and surface downward shortwave radiation and weekly wastewater surveillance data on levels of three respiratory viruses as covariates. We employed a Monte Carlo simulation approach, performing 1,000 model iterations to estimate 95% empirical confidence intervals (eCIs) for the predictions.

**eTable 7. Estimated change in the percent of virtual acute care visits for all-cause, cardiovascular, injury, neuropsychiatric, and respiratory endpoints among Kaiser Permanente Southern California (KPSC) highly-exposed members (n=305,258) in the two weeks following the January 7, 2025 ignition of the LA Fires.**

| *Percent (95% eCI)* | | | | | | |
|---|---|---|---|---|---|---|
| **Date** | **Weekday** | **All-cause** | **Cardiovascular** | **Injury** | **Neuropsychiatric** | **Respiratory** |
| 2025-01-07 | Tuesday | 14% (-9%, 36%) | 30% (7%, 57%) | 18% (2%, 36%) | 12% (-4%, 28%) | 35% (23%, 47%) |
| 2025-01-08 | Wednesday | 5% (-17%, 31%) | 32% (14%, 58%) | -19% (-47%, 3%) | -8% (-26%, 14%) | 36% (23%, 50%) |
| 2025-01-09 | Thursday | 15% (-7%, 37%) | 36% (18%, 61%) | 8% (-12%, 25%) | 8% (-8%, 25%) | 43% (32%, 54%) |
| 2025-01-10 | Friday | 20% (-5%, 45%) | 44% (29%, 66%) | -4% (-30%, 17%) | 8% (-14%, 31%) | 47% (35%, 58%) |
| 2025-01-11 | Saturday | 58% (-59%, 183%) | -70% (-364%, 308%) | 12% (-83%, 89%) | 4% (-85%, 101%) | 50% (0%, 96%) |
| 2025-01-12 | Sunday | 95% (-102%, 324%) | -114% (-591%, 585%) | -294% (-807%, 147%) | 17% (-153%, 207%) | 49% (1%, 91%) |
| 2025-01-13 | Monday | 24% (4%, 49%) | 34% (16%, 63%) | 24% (5%, 42%) | 18% (2%, 35%) | 39% (27%, 50%) |
| 2025-01-14 | Tuesday | 16% (-3%, 38%) | 30% (12%, 56%) | 11% (-6%, 28%) | 12% (-2%, 28%) | 25% (12%, 36%) |
| 2025-01-15 | Wednesday | 15% (-5%, 38%) | 31% (13%, 57%) | 11% (-9%, 29%) | 12% (-4%, 28%) | 39% (27%, 49%) |
| 2025-01-16 | Thursday | 26% (6%, 54%) | 41% (25%, 68%) | 6% (-13%, 27%) | 21% (5%, 40%) | 36% (23%, 47%) |
| 2025-01-17 | Friday | 19% (-6%, 44%) | 25% (4%, 55%) | -30% (-60%, -3%) | 16% (-3%, 37%) | 30% (16%, 43%) |
| 2025-01-18 | Saturday | 53% (-81%, 189%) | -308% (-1009%, 706%) | -24% (-178%, 108%) | 20% (-58%, 103%) | 22% (-43%, 85%) |
| 2025-01-19 | Sunday | 98% (-109%, 309%) | -154% (-741%, 703%) | 37% (-72%, 129%) | 20% (-154%, 203%) | 45% (-15%, 108%) |
| 2025-01-20 | Monday | -119% (-186%, -35%) | -3462% (-4562%, -1734%) | -2047% (-2609%, -1447%) | -37% (-66%, 1%) | -143% (-195%, -91%) |

We used the maximum wildfire burn zone reached by an LA or Ventura County wildfire as of January 16, 2025 to define exposure. Highly exposed members resided in a census tract located <20km burn zones. Results from an interrupted time series model using KPSC electronic health record data from November to January 2022–2025, with daily maximum and minimum temperature and humidity, wind velocity, and surface downward shortwave radiation and weekly wastewater surveillance data on levels of three respiratory viruses as covariates. We employed a Monte Carlo simulation approach, performing 1,000 model iterations to estimate 95% empirical confidence intervals (eCIs) for the predictions.

**eTable 8. Estimated change in the percent of virtual acute care visits for all-cause, cardiovascular, injury, neuropsychiatric, and respiratory endpoints among Kaiser Permanente Southern California (KPSC) moderately-exposed members (n=1,373,419) in the two weeks following the January 7, 2025 ignition of the LA Fires.**

| *Percent (95% eCI)* | | | | | | |
|---|---|---|---|---|---|---|
| **Date** | **Weekday** | **All-cause** | **Cardiovascular** | **Injury** | **Neuropsychiatric** | **Respiratory** |
| 2025-01-07 | Tuesday | 10% (-8%, 27%) | 31% (9%, 58%) | 16% (2%, 30%) | 14% (-7%, 33%) | 29% (13%, 44%) |
| 2025-01-08 | Wednesday | 14% (-4%, 32%) | 38% (18%, 64%) | 9% (-8%, 26%) | 12% (-8%, 33%) | 35% (23%, 50%) |
| 2025-01-09 | Thursday | 19% (3%, 37%) | 35% (15%, 61%) | 11% (-6%, 28%) | 19% (0%, 39%) | 36% (22%, 51%) |
| 2025-01-10 | Friday | 21% (1%, 43%) | 39% (18%, 67%) | 21% (5%, 38%) | 23% (1%, 49%) | 37% (23%, 53%) |
| 2025-01-11 | Saturday | 58% (-29%, 151%) | 3% (-191%, 248%) | 59% (-16%, 137%) | 27% (-58%, 120%) | 31% (-18%, 83%) |
| 2025-01-12 | Sunday | 94% (-40%, 265%) | -23% (-520%, 704%) | 60% (-67%, 209%) | 44% (-131%, 248%) | 25% (-22%, 75%) |
| 2025-01-13 | Monday | 27% (10%, 47%) | 36% (14%, 66%) | 26% (10%, 45%) | 25% (4%, 48%) | 37% (25%, 50%) |
| 2025-01-14 | Tuesday | 14% (-3%, 34%) | 33% (14%, 59%) | 6% (-10%, 25%) | 14% (-3%, 35%) | 27% (14%, 41%) |
| 2025-01-15 | Wednesday | 16% (-2%, 35%) | 28% (8%, 58%) | 10% (-6%, 28%) | 14% (-6%, 36%) | 27% (13%, 42%) |
| 2025-01-16 | Thursday | 25% (8%, 47%) | 34% (13%, 63%) | 10% (-7%, 34%) | 22% (3%, 44%) | 36% (23%, 48%) |
| 2025-01-17 | Friday | 19% (0%, 42%) | 25% (1%, 56%) | 14% (-4%, 33%) | 18% (-8%, 46%) | 28% (12%, 42%) |
| 2025-01-18 | Saturday | 54% (-39%, 155%) | -32% (-320%, 389%) | 21% (-135%, 188%) | 24% (-77%, 135%) | 21% (-27%, 67%) |
| 2025-01-19 | Sunday | 80% (-72%, 242%) | -47% (-438%, 492%) | 56% (-77%, 199%) | 36% (-159%, 241%) | 37% (-15%, 90%) |
| 2025-01-20 | Monday | -130% (-183%, -63%) | -1221% (-1679%, -620%) | -289% (-377%, -183%) | -48% (-90%, -8%) | -54% (-90%, -18%) |

We used the maximum wildfire burn zone reached by an LA or Ventura County wildfire as of January 16, 2025 to define exposure. Moderately exposed members lived in tracts ≥20km from a burn zone but within LA County. Results from an interrupted time series model using KPSC electronic health record data from November to January 2022–2025, with daily maximum and minimum temperature and humidity, wind velocity, and surface downward shortwave radiation and weekly wastewater surveillance data on levels of three respiratory viruses as covariates. We employed a Monte Carlo simulation approach, performing 1,000 model iterations to estimate 95% empirical confidence intervals (eCIs) for the predictions.

**eTable 9. Traditional difference-in-differences based estimation of change in the number of (A) outpatient and (B) virtual acute care visits for all-cause, cardiovascular, injury, neuropsychiatric, and respiratory endpoints at Kaiser Permanente Southern California (KPSC) in the week following the January 7, 2025 ignition of the 2025 LA Fires.**

| Estimated excess encounters | | | | | | |
|---|---|---|---|---|---|---|
| **Exposure group** | **Encounter type** | **All-cause** | **Cardiovascular** | **Injury** | **Neuropsychiatric** | **Respiratory** |
| Highly | Outpatient | -3002 | -305 | -117 | -480 | -3 |
| Highly | Virtual | 597 | 212 | -11 | 328 | 235 |
| Moderately | Outpatient | 694 | 1812 | -169 | 452 | 929 |
| Moderately | Virtual | 5425 | 596 | 73 | 3028 | 1219 |

Based on the maximum wildfire burn zone reached by an LA or Ventura County wildfire as of January 16, 2025, highly exposed members resided in a census tract located <20km from a burn zone and moderately exposed members lived in tracts ≥20km but within LA County.
